# Local causal discovery in epidemiology: an application to quantifying the effect of diabetes on severe liver fibrosis in patients with viral hepatitis

**DOI:** 10.1101/2025.09.02.25334768

**Authors:** Timothée Loranchet, Daria Bystrova, Paul Burgat, Jonathan Bellet, Marc Bourlière, Clovis Lusivika-Nzinga, Jerome Nicol, Lucia Parlati, Pierre-Yves Boëlle, Fabrice Carrat, Charles K. Assaad

## Abstract

**Background:** Estimating the controlled direct effect (CDE) from observational data is challenging when the DAG is unknown. Causal discovery methods can infer a partially oriented DAG, enabling the identification of potential adjustment sets. We use a local causal discovery algorithm that focuses on the relevant portion of the graph, reducing assumptions and complexity compared to global methods. This approach is applied to a viral hepatitis cohort to estimate the CDE of diabetes on severe liver fibrosis.

**Methods:** The CDE of diabetes on liver fibrosis in patients with HBV or HCV was assessed using baseline data from the French ANRS CO22 HEPATHER cohort initiated in 2012. A local causal discovery algorithm, LocalPC-CDE, with bootstrap augmentation identified a robust adjustment set, retaining only variables minimally affected by sampling variability. The CDE was quantified as a causal odds ratio using logistic regression.

**Results:** Causal discovery included 20858 patients, with estimation performed on 8802 complete-case observations. The algorithm identified an adjustment set of seven variables: geographical origin, age, hepatitis type, total cholesterol, HDL cholesterol, past alcohol consumption, blood glucose, and sex. The CDE of diabetes on severe fibrosis in viral hepatitis patients was significantly positive, with an estimated odds ratio of 2.03 (95% CI [1.78, 2.31]).

**Conclusions:** After causal adjustment using a targeted, data-driven approach, diabetes retained a direct and statistically significant effect on liver fibrosis in patients with chronic viral hepatitis. This paper more generally introduces a methodological pipeline for local causal discovery when the underlying DAG is uncertain.

## 1 INTRODUCTION

Randomized trials are often infeasible or unethical in epidemiology, making observational data the primary resource for causal investigation. To draw valid conclusions from such data, causal inference frameworks such as structural causal models (SCMs) and directed acyclic graphs (DAGs) provide formal tools for estimating causal effects under explicit assumptions [1, 2, 3]. Of particular interest is the *controlled direct effect* (CDE) [3, 2], which captures the effect of an exposure on an outcome that is not transmitted through mediating variables. When the underlying DAG is unknown, *causal discovery* methods can help infer it directly from the data [4, 5, 6, 7, 8]. Unlike classical approaches that aim to recover the entire DAG, *local* causal discovery restricts attention to the part of the graph relevant for a given question. This reduces assumptions, improves computational efficiency, and yields valid adjustment sets for causal effect estimation, including the CDE.

In this study, we apply the local causal discovery algorithm LocalPC-CDE [9] to baseline data from the French *ANRS CO22 HEPATHER* cohort, which collects detailed clinical, biological, and virological information on patients with chronic viral hepatitis. Chronic infection with hepatitis B (HBV) or hepatitis C (HCV) is a major cause of liver-related morbidity, due to the risk of progression to cirrhosis, hepatocellular carcinoma, and liver failure [10, 11, 12, 13]. Liver fibrosis, the accumulation of extracellular matrix proteins in response to chronic injury, is a key determinant of prognosis and is staged from F0 (no fibrosis) to F4 (cirrhosis), with severe fibrosis defined as stage F3–F4.

Diabetes mellitus is a chronic metabolic disorder characterized by hyperglycemia due to impaired insulin secretion or action. Observational studies have reported associations between diabetes and advanced fibrosis [10, 11, 12, 13, 14, 15, 16], but the quantification of the diabetes *direct causal effect* on liver fibrosis, independently of other mediating factors, has received little attention. Using local causal discovery, combined with robustness checks such as bootstrap augmentation [17] and sensitivity analyses [18], we estimate the CDE of diabetes on severe fibrosis, showing that indeed diabetes exerts directly affects liver fibrosis.

Beyond this specific application to HEPATHER, the broader aim of this paper is to illustrate how local causal discovery can complement, rather than replace, traditional epidemiological approaches for causal analysis. By focusing only on the parts of the DAG relevant to a given question, local discovery provides a practical way to identify valid adjustment sets and quantify causal effects, especially when the full DAG is uncertain. To make our approach more transparent, Figure 1 presents a pipeline that visually summarizes the steps we followed to estimate the controlled direct effect of diabetes on severe fibrosis in the HEPATHER cohort.

**Figure 1.**
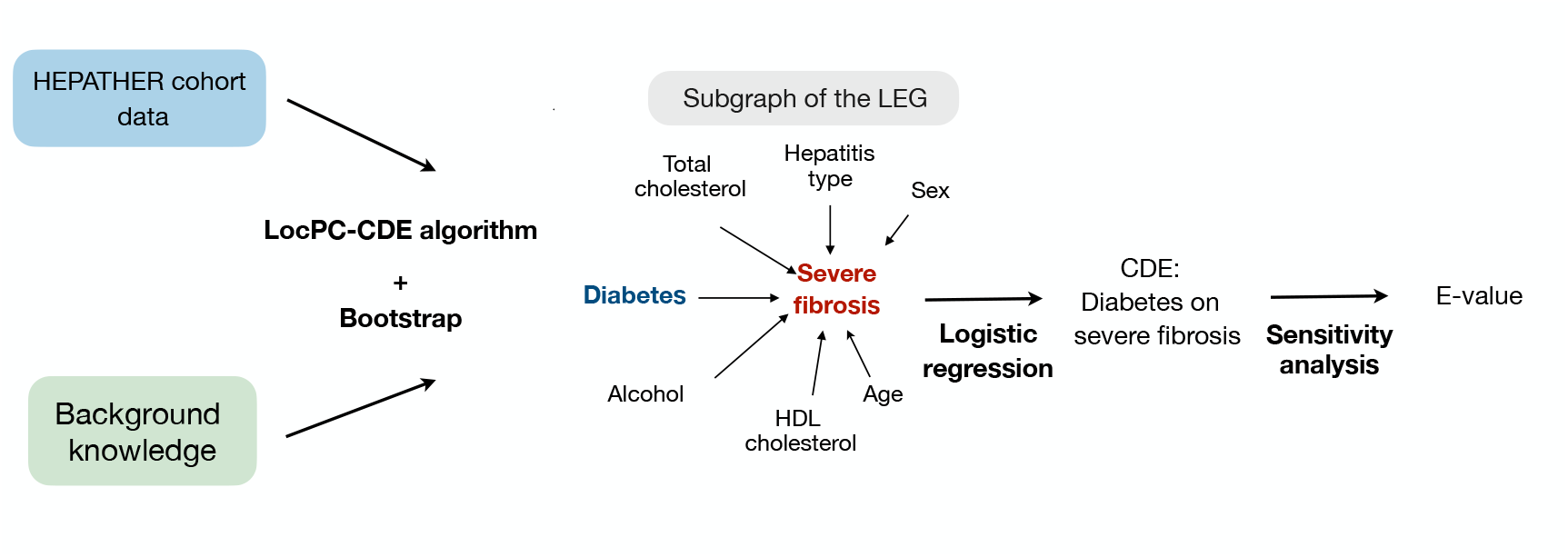
Schematic representation of the procedure for estimating the CDE of diabetes in severe fibrosis using LocalPC-CDE algorithm.

## 2 MATERIEL AND METHODS

### 2.1 Controlled Direct Effect

Let *D* ∈ {0, 1}denote the diabetes status (1 = diabetic, 0 = healthy), *F ∈* {0, 1}the severe fibrosis (1 = stage ≥ F3, 0 = stage *<* F3), and **Z** the set of the observed direct causes of fibrosis (except diabete) in the causal DAG.

A *mediator* is a variable that lies on the causal path from diabetes to fibrosis (e.g., a variable *M* such that *D → M → F*). If such mediators exist, our goal is to estimate the effect of diabetes on fibrosis that does *not* operate through them, this is referred to as the *direct* effect along the path *D → F*. One way to formalize and estimate this quantity is through the *Controlled Direct Effect* (CDE) [3, 2]. The CDE represents the effect of setting diabetes to a fixed value while holding all other direct causes of fibrosis at specific values. From a causal perspective, this corresponds conceptually to an intervention where diabetes is set to *D* = 1 (denoted *do*(*D* = 1)), and all other direct causes of fibrosis are fixed to specific values **Z** = **z** via *do*(**Z** = **z**). Then, we compare this to a scenario where diabetes is set to *D* = 0 while keeping the same intervention on **Z**. The CDE of diabetes (*D*) on severe fibrosis (*F*) can then be quantified using an odds ratio comparing the two interventional distributions:

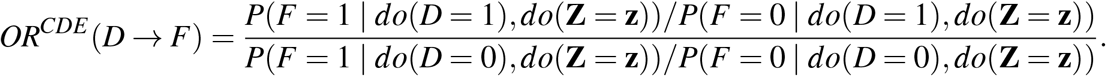

To estimate this causal effect from observational data, three patterns must be considered: (1) confounders (variables causing both diabetes and severe fibrosis), which may bias the estimation if not adjusted for; (2) mediators (intermediate variables caused by diabetes and in turn causing fibrosis), which must be adjusted for to estimate the CDE; and (3) colliders [1] (variables caused by both diabetes and fibrosis), which should not be adjusted for, as doing so would introduce collider bias [19, 20]. Conditioning on all direct causes of fibrosis simultaneously blocks bias due to confounders and influence of mediators, while preventing adjustment for colliders. Under the assumption of no unmeasured confounding, the set of direct causes of fibrosis **Z** thus constitutes a causally valid adjustment set, in the sense that the causal odds ratio *OR*^*CDE*^ (*D → F*) can be identified from observational data. This identifiability is expressed by the equivalence between interventional and conditional probabilities:

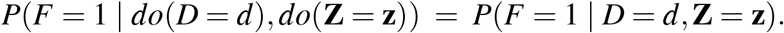

Remark that **Z** is not only valid but also optimal in the sense that it minimizes the asymptotic estimation variance [21]. To estimate the CDE by adjusting on **Z** we use a logistic regression model, with 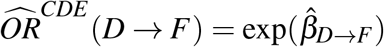. The reported 95% confidence intervals are computed as 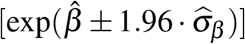, where 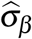 denotes the estimated standard error of the coefficient 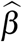 . The models are fitted on the complete cases.

To assess the sensitivity of 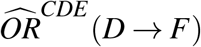 to potential unmeasured confounding between diabetes and severe fibrosis, we compute the associated *E-value* [18]. The E-value quantifies the minimum strength of association that an unmeasured confounder would need to have with both the exposure and the outcome, conditional on the measured covariates, in order to fully explain away this estimated effect.

The main difficulty thus lies in identifying such a set of variables **Z**. The use of Directed Acyclic Graphs (DAGs) is a powerful tool for this purpose: the required variables are simply those with a directed edge pointing to *F* in the DAG. If the DAG is known (e.g., based on expert knowledge), then this identification is straightforward. However, the DAG may also be partially or completely unknown. In such cases, (local) causal discovery becomes a useful approach.

### 2.2 Local Causal Discovery Procedure

We assume here that the DAG is not fully known a priori and must be inferred from the data. This task, known as *causal discovery*, is both computationally and theoretically challenging. Recent approaches suggest that instead of recovering the entire DAG, it is often sufficient to estimate only a *local* portion of the graph surrounding the target (called a Local Essential Graph, LEG) [9]. Such local structures are enough to identify the direct causes of the target variable, and they can be learned more efficiently and robustly. This targeted approach is referred to as *local causal discovery*. Figure 2 illustrates the difference between global and local causal discovery.

**Figure 2.**
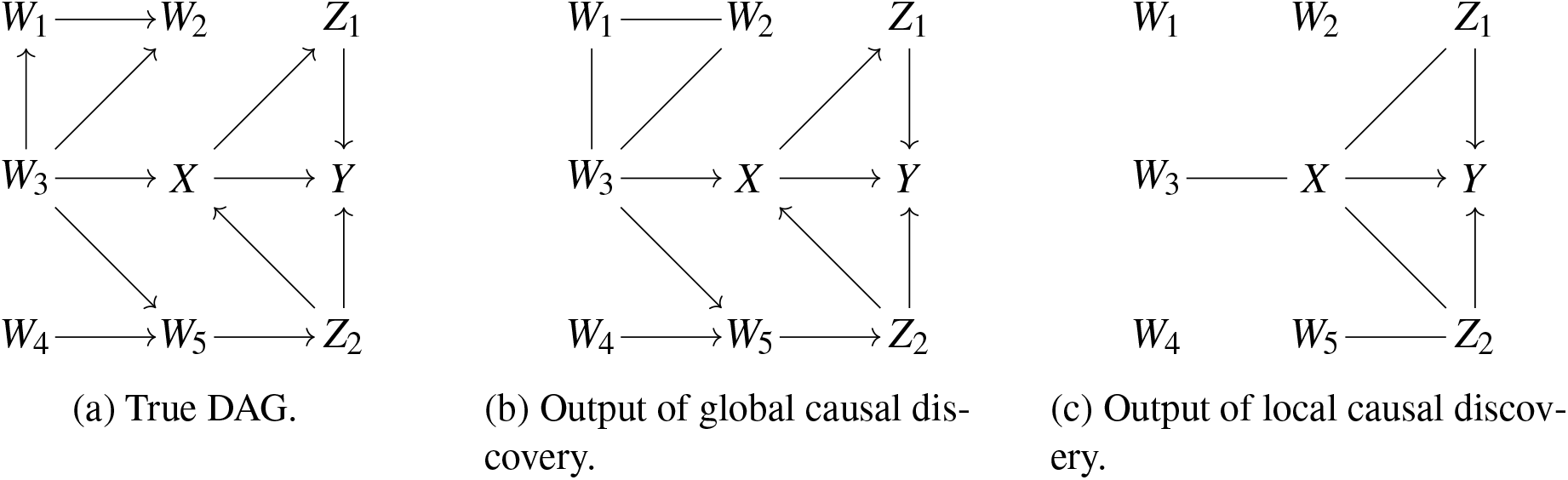
Illustration of the difference between global and local causal discovery. (a) The underlying true DAG. (b) The output of PC, a global causal discovery algorithm, which attempts to recover the entire graph and in this example requires around 490 conditional independence tests. (c) The output of LocPC-CDE, a local causal discovery algorithm focused only on the part of the graph relevant for estimating a CDE, requiring only about 250 tests.

#### Local Causal Discovery Algorithm

Local causal discovery is performed with severe fibrosis as the target variable by using the LocPC-CDE algorithm [9] for causal discovery. We briefly describe here the functioning of the LocPC-CDE algorithm; for a detailed explanation and theoretical guarantees, see [9]. This method relies on conditional independence testing, following principles similar to the well-known Peter-Clark (PC) algorithm [4] introduced for global causal discovery (see [22, 23, 24, 25] for applications of the global PC algorithm to epidemiological data). LocPC-CDE begins by testing for conditional independencies between the target variable (severe fibrosis) and all other variables to identify its direct neighbors. Then, edge orientations are inferred using conditional independence information combined with logical orientation rules (Meek’s rules [26]). The goal is to orient all edges adjacent to the target. The more of the graph is discovered, the more orientations become possible. To facilitate this, local discovery is iteratively extended to nodes adjacent to the target, then to their neighbors, and so on, progressively revealing and orienting the *neighborhood* of the target. The algorithm stops automatically (and optimally, in the sense that it minimizes the neighborhood size and thus the number of tests and potential errors) once all edges adjacent to the target are oriented (if this condition is never met, the algorithm returns that the CDE is not identifiable). The algorithm requires an appropriate conditional independence test. To ensure reasonable computation time and a consistent testing procedure, we choose to discretize continuous variables in order to apply a parametric independence test. Discretization is performed using clinical thresholds [27] as detailed in Table 1. Conditional independence is then assessed using the *G*^2^ test (likelihood ratio test for categorical variables [28]). The LocPC-CDE algorithm is consistent in recovering the correct LEG under the assumptions of causal sufficiency (no unmeasured confounder) and local faithfulness, a standard assumption stating that observed conditional independencies reflect the underlying causal structure in the variables’ local neighborhood.

**Table 1:**
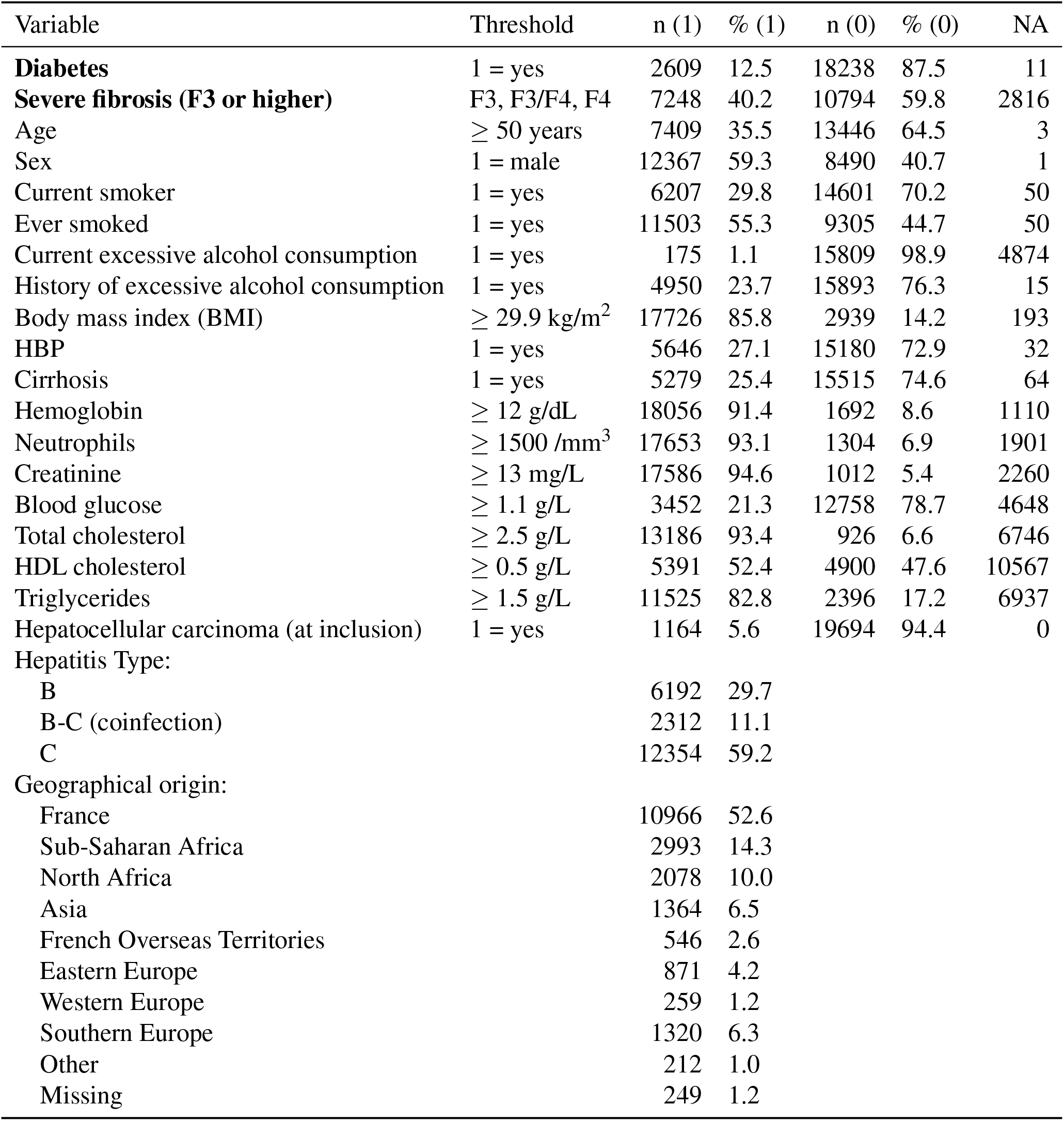
Variables on which local causal discovery is performed (*N* = 20858).

| Variable | Threshold | n (1) | % (1) | n (0) | % (0) | NA |
| --- | --- | --- | --- | --- | --- | --- |
| **Diabetes** | 1 = yes | 2609 | 12.5 | 18238 | 87.5 | 11 |
| **Severe fibrosis (F3 or higher)** | F3, F3/F4, F4 | 7248 | 40.2 | 10794 | 59.8 | 2816 |
| Age | $\geq 50$ years | 7409 | 35.5 | 13446 | 64.5 | 3 |
| Sex | 1 = male | 12367 | 59.3 | 8490 | 40.7 | 1 |
| Current smoker | 1 = yes | 6207 | 29.8 | 14601 | 70.2 | 50 |
| Ever smoked | 1 = yes | 11503 | 55.3 | 9305 | 44.7 | 50 |
| Current excessive alcohol consumption | 1 = yes | 175 | 1.1 | 15809 | 98.9 | 4874 |
| History of excessive alcohol consumption | 1 = yes | 4950 | 23.7 | 15893 | 76.3 | 15 |
| Body mass index (BMI) | $\geq 29.9$ kg/m<sup>2</sup> | 17726 | 85.8 | 2939 | 14.2 | 193 |
| HBP | 1 = yes | 5646 | 27.1 | 15180 | 72.9 | 32 |
| Cirrhosis | 1 = yes | 5279 | 25.4 | 15515 | 74.6 | 64 |
| Hemoglobin | $\geq 12$ g/dL | 18056 | 91.4 | 1692 | 8.6 | 1110 |
| Neutrophils | $\geq 1500$ /mm<sup>3</sup> | 17653 | 93.1 | 1304 | 6.9 | 1901 |
| Creatinine | $\geq 13$ mg/L | 17586 | 94.6 | 1012 | 5.4 | 2260 |
| Blood glucose | $\geq 1.1$ g/L | 3452 | 21.3 | 12758 | 78.7 | 4648 |
| Total cholesterol | $\geq 2.5$ g/L | 13186 | 93.4 | 926 | 6.6 | 6746 |
| HDL cholesterol | $\geq 0.5$ g/L | 5391 | 52.4 | 4900 | 47.6 | 10567 |
| Triglycerides | $\geq 1.5$ g/L | 11525 | 82.8 | 2396 | 17.2 | 6937 |
| Hepatocellular carcinoma (at inclusion) | 1 = yes | 1164 | 5.6 | 19694 | 94.4 | 0 |
| Hepatitis Type: |  |  |  |  |  |  |
| B |  | 6192 | 29.7 |  |  |  |
| B-C (coinfection) |  | 2312 | 11.1 |  |  |  |
| C |  | 12354 | 59.2 |  |  |  |
| Geographical origin: |  |  |  |  |  |  |
| France |  | 10966 | 52.6 |  |  |  |
| Sub-Saharan Africa |  | 2993 | 14.3 |  |  |  |
| North Africa |  | 2078 | 10.0 |  |  |  |
| Asia |  | 1364 | 6.5 |  |  |  |
| French Overseas Territories |  | 546 | 2.6 |  |  |  |
| Eastern Europe |  | 871 | 4.2 |  |  |  |
| Western Europe |  | 259 | 1.2 |  |  |  |
| Southern Europe |  | 1320 | 6.3 |  |  |  |
| Other |  | 212 | 1.0 |  |  |  |
| Missing |  | 249 | 1.2 |  |  |  |

#### Significance Threshold

As previously discussed, the LocPC-CDE algorithm relies on conditional independence tests to infer the LEG in the vicinity of the target variable. This requires specifying a significance threshold *α*, which determines the rejection criterion for the null hypothesis of conditional independence. The significance threshold *α* does not represent a significance level for the estimated graph, as the LocPC-CDE algorithm conduct a sequence of conditional independence tests, where the outcome of each test depends on the results of the previous tests. However, *α* can be considered as a sparsity level: a larger value of *α* tends to produce a denser estimated local graph, with more edges retained, while a smaller *α* yields a sparser graph, with fewer edges included. In our analysis, we adopt the conventional threshold of *α* = 0.05.

#### Background knowledge

Causal discovery algorithms can generally incorporate background knowledge, that is, prior information on certain causal relationships (e.g., derived from expert opinion, the literature, or previous studies). Such knowledge can take different forms, but in this work we focus on orientation constraints: we pre-specify that some edges, if present, must be oriented in a certain direction. Importantly, this does not force the algorithm to include the edge, but rather ensures that if it is detected, its orientation is consistent with prior knowledge. Incorporating background knowledge in this way can reduce computational time and improve algorithmic performance, provided that the prior information is correct. Moreover, these constraints do not need to concern only liver fibrosis: any reliable orientation can improve the learning process. In this application, we impose some of such constraints. First, no variable in the model can cause age, sex, or geographical origin; therefore, if an edge connects to these variables, it cannot be oriented towards them (e.g., liver fibrosis cannot cause age). Second, diabetes is constrained to be a cause of severe fibrosis, given consistent evidence from both epidemiological data and biological mechanisms. Third, by definition, cirrhosis is a consequence of severe fibrosis, and hepatocellular carcinoma is a consequence of fibrosis (and potentially cirrhosis); thus, any edges among these variables must point from fibrosis towards cirrhosis or carcinoma. Finally, for viral hepatitis, we specify that the type of infection is viral and then is not caused by the measured variables of the model, except for potential causes from geographical origin (e.g., higher prevalence in specific regions[29]), sex (e.g., behavioral risk factors[30]), and age[29].

#### Missing data

The local causal discovery algorithms we use assume complete data for correct operation, and missing values can potentially introduce bias, especially if the missingness is related to other variables. To handle this, we assume that the data are *Missing Completely At Random* (MCAR) and apply a *test-wise deletion* strategy [31]. That is, for each statistical test, we use only individuals with observed values for the variables involved in that test, while keeping individuals with missing values in unrelated variables. For instance, when testing the conditional independence of *X* and *Y* given **Z**, we exclude rows with missing values in *X, Y*, or any variable in **Z**, but retain rows with missing values in other variables. Under the MCAR assumption, this approach preserves the validity of the discovery algorithms and maximizes the statistical power of each test.

#### Uncertainty on the inferred causal structure

Running the LocPC-CDE algorithm leads to a point estimate of the LEG, which is subject to finite-sample bias. Such bias may affect conditional independence tests, potentially leading to spurious edges, missing true edges, or incorrect edge orientations. The main issue is the absence of an uncertainty measure: would a slightly different sample have led to the same result? To address this, a bootstrap step is added into the discovery process, as suggested in [17]. We draw *B* bootstrap samples of size *N*, run the local causal discovery algorithm on each sample, and measure the frequency of a feature of interest across the *B* outputs. We apply this procedure exclusively in our real-data application. In this context, when estimating the CDE of diabetes on severe fibrosis, our interest lies in identifying the set of direct causes of severe fibrosis (denoted *F*). We therefore assess, for a given *N*, the confidence that *Z* is a direct cause of *F* as:

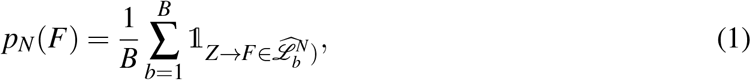

where 𝟙 is the indicator function and 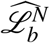 is the *b*-th inferred LEG from sample of size *N*. Since the algorithm is consistent for recovering the LEG, this quantity should converge to 1 for true direct causes of fibrosis, and to 0 for non-causes as *N* increases. We use the full dataset size in the bootstrap procedure *N* = 20858, and set *B* = 100. Variables are selected as direct causes of severe fibrosis if they are identified in more than 50% of the bootstrap samples.

### 2.3 Data

Data were obtained from the ANRS CO22 HEPATHER cohort, which is a French national, multicenter, prospective, observational cohort study of patients with hepatitis B infection or hepatitis C (past or present) infection [32]. Initiated in 2012, this prospective cohort follows participants for approximately ten years to improve understanding and management of new hepatitis treatments and currently includes 20858 patients, see details in [32]. In this study, we used sociodemographic, clinical, and biological data collected at the time of cohort enrollment from patients with chronic or resolved hepatitis B or C.

#### Variables

We selected 21 baseline variables potentially related to liver fibrosis: (1) the outcome *severe fibrosis* (fibrosis stage F3 or higher); (2) the exposure *diabetes* (diagnosis of diabetes); (3) other variables *age* (years at inclusion), *sex* (biological sex), *current smoker* and *ever smoked* (tobacco use status), *current* and *history of excessive alcohol consumption, body mass index* (BMI, weight-to-height ratio), *HBP* (history of high blood pressure), *cirrhosis* (most advanced stage of liver fibrosis), *hemoglobin* (blood oxygen-carrying capacity), *neutrophils* (immune cell count), *creatinine* (renal function marker), *blood glucose* (glycemia), *total cholesterol* (overall blood cholesterol concentration), *HDL cholesterol* (“good” cholesterol fraction), *triglycerides* (blood lipid level), *hepatocellular carcinoma at inclusion* (HCC, liver cancer status), *hepatitis type* (B,C or BC), and *geographical origin* (region of birth).

We excluded liver fibrosis biomarkers (e.g., gammagt, albumin, alat, etc.) to avoid deterministic or near-deterministic relations that could bias causal discovery, as they would violate the local faithfulness assumption and might prevent detecting causal relationships (if a variable (the proxy) is a quasi-deterministic function of fibrosis, it renders nearly all other variables conditionally independent of it, since almost all information about fibrosis is contained in this proxy).

#### Models fitting

The local causal discovery algorithm with bootstrap is then applied to this sample to identify the direct causes of severe fibrosis the causal odd ratio is estimated. This model, adjusted on a causally valid set, is compared to two “non-causal” logistic models: (1) a naive model, adjusting for no covariates and assessing the univariate association between diabetes and severe fibrosis; and (2) an overadjusted model, controlling for all variables in the dataset. Each model is fitted on the complete data corresponding to its included variables. Furthermore, although local causal discovery is performed on discretized data, the original continuous measurements are available. To strengthen the robustness of our findings, a logistic regression model including the continuous variables alongside the model derived from the discretized dataset is fitted.

We used Python 3.13.3 for LocPC-CDE and R 4.5.0 (glm, EValue) for logistic regression and E-value computation.

## 3 RESULTS

### 3.1 Estimation of direct causes

The bootstrap procedure output is shown in Figure 3, which displays the proportion of bootstrap samples in which each variable is identified as a direct cause of severe fibrosis, along with the corresponding 95% bootstrap confidence intervals. We retain all these variables as direct causes, as they are identified as such in over 50% of the samples: age, hepatitis type, HDL cholesterol, geographical origin, sex, total cholesterol, and alcohol history.

**Figure 3.**
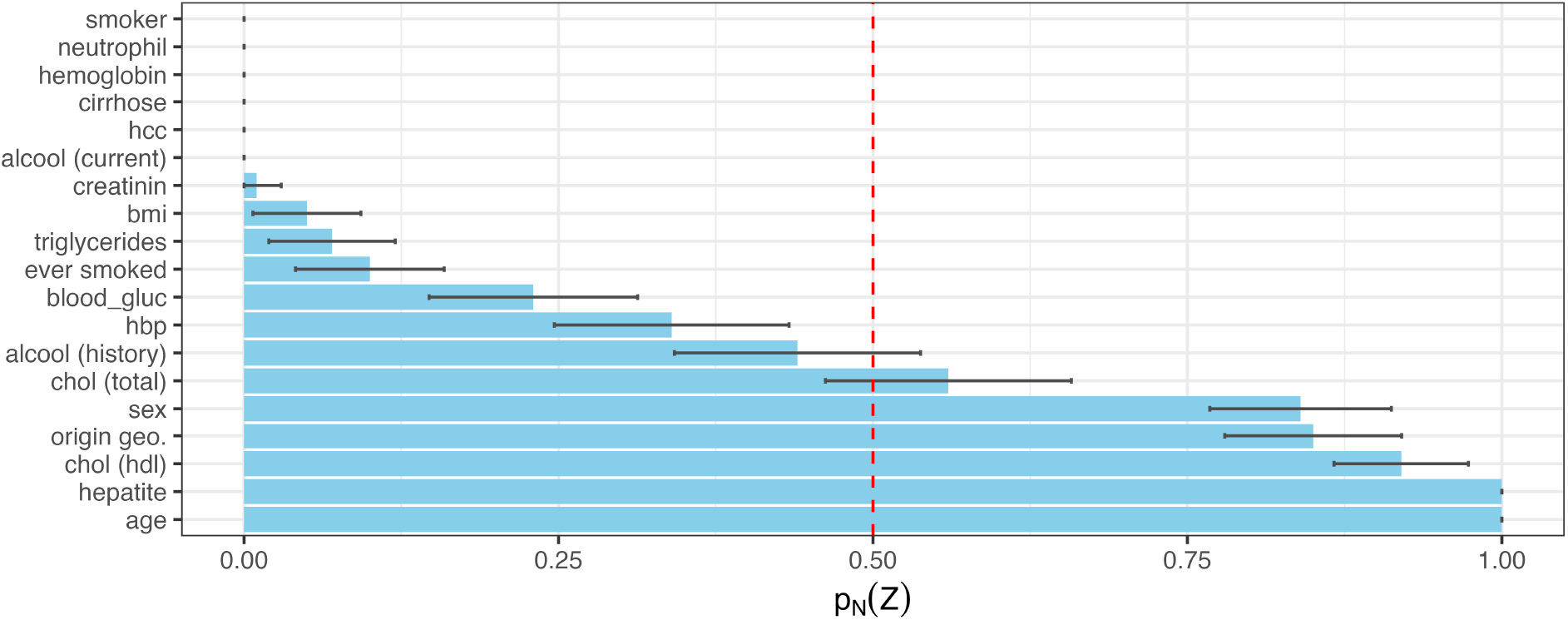
Confidence *p*_*N*_(*Z*) that each variable *Z* is a direct cause of severe fibrosis with 95% bootstrap CI intervals. Red = 50%. *B* = 100.

Other variables are rarely or never identified as direct causes of severe fibrosis: current smoking, alcohol consumption, neutrophils, hemoglobin, cirrhosis, and hepatocarcinoma are never direct causes; creatinine, BMI, triglycerides, and history of smoking are only rarely identified; while blood glucose and hypertension are occasionally flagged. We attribute these occasional assignments to finite-sample noise and the algorithm’s sensitivity to minor errors that can induce incorrect edge orientations. Notably, our method only constrains edge orientations based on background knowledge, not the presence of edges themselves. For example, age is identified as a direct cause of fibrosis in 100% of bootstrap samples, indicating that no sample showed a conditional independence between age and severe fibrosis, consistent with a robust causal link. These findings are consistent with the existing literature: age, past alcohol consumption, and sex have been associated with severe fibrosis in patients with viral hepatitis [33] and low HDL cholesterol is a known risk factor for liver disease [34]. The absence of a direct link is consistent: blood glucose, showing a weak signal here, has been reported to have no significant effect on severe fibrosis independent of diabetes [35], confirming its non-direct cause status.

Thus, the set of variables **Z** ={age, geographic origin, hepatitis type, sex, total and HDL cholesterol, alcohol consumption history} forms our presumed causally valid adjustment set for the logistic model.

### 3.2 CDE estimation

The model adjusted on the estimated direct causes of severe fibrosis is presented in Table 2. The causal odds ratio corresponding to the CDE of diabetes on severe fibrosis is **2.03 (95% CI [1.78; 2.31])**, indicating that, among patients with hepatitis B or C, having diabetes *causes* a 2.03-fold increase (95% CI [1.78; 2.31]) in the odds of severe fibrosis.

**Table 2:**
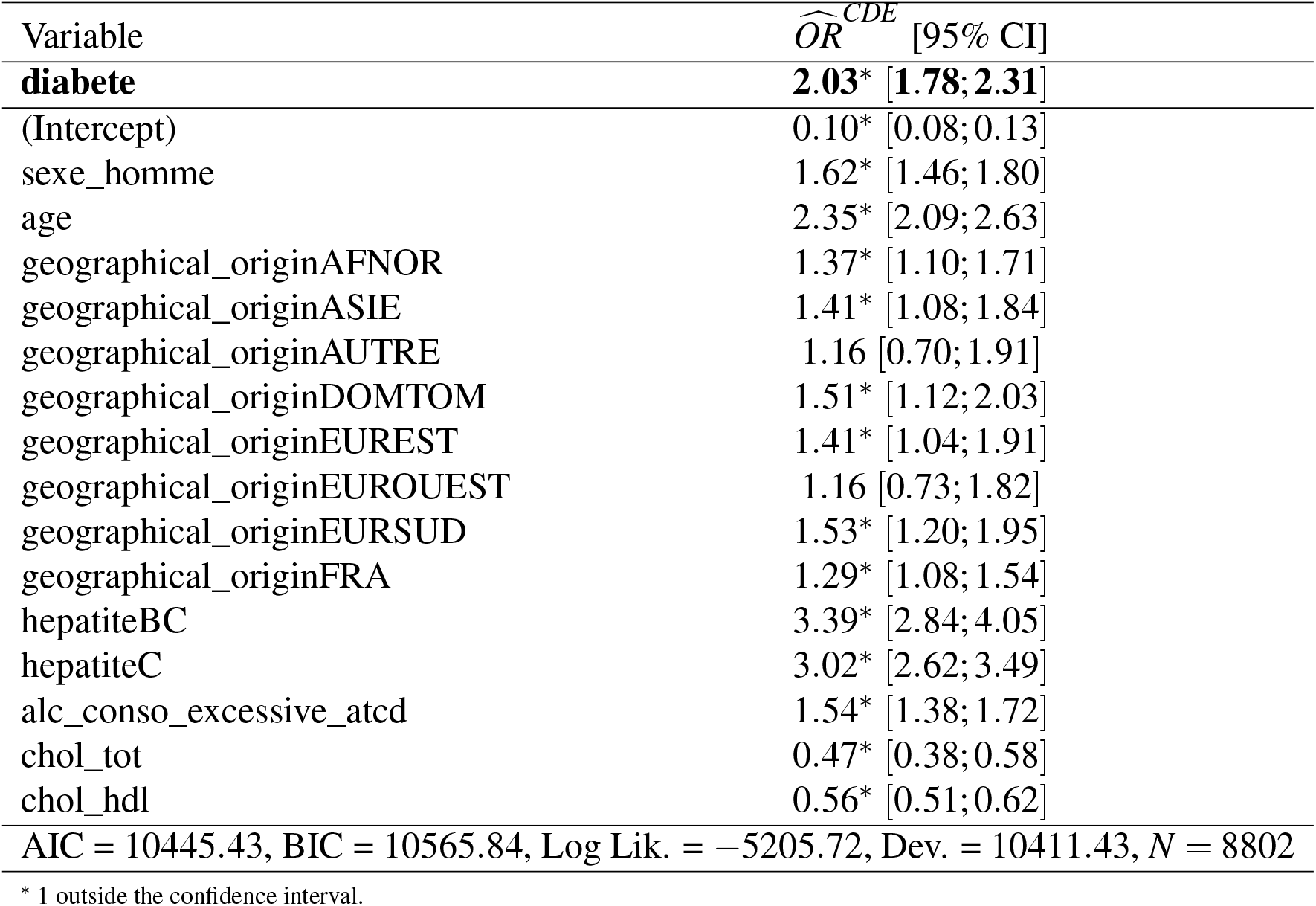
Logistic Regression with Odds Ratios.

While this addresses the primary question of this study, it is noteworthy that the adjustment set would be the same to estimate the CDE of any other variable on severe fibrosis, as the direct causes of fibrosis constitute a set that is independent of the exposure under study. Consequently, all odds ratios in Table 2 can be interpreted causally as CDE odds ratios. In this particular case (since we are studying CDEs), the table avoids the Table 2 Fallacy [36], which typically cautions that coefficients other than the exposure of interest in a multivariable regression cannot be interpreted causally.

The sensitivity analysis using the E-value [18] yielded an estimate of 4.85 (lower CI bound of 4.01) expressed in odds ratios [37]. This implies that, to reduce the observed odds ratio for the effect of diabetes on severe fibrosis to be non-significant, an unmeasured confounder would need to be associated with both diabetes and severe fibrosis with an odds ratio of at least 4.01 each. The E-value does not define a robustness threshold; its interpretation relies on expert judgment regarding the plausibility of such unmeasured confounding. In our context, such an effect size ranging from 4.01 to 4.85 would be among the largest odds ratios observed in our logistic regression models, comparable in magnitude to the effect of hepatitis type, suggesting that only a really strong unmeasured confounder could fully explain away the positive association.

Finally, Figure 4 compares the causal discovery based estimation with two other models (no adjustment and adjustment on all variables). The naive simple regression model substantially overestimates the odds ratio of diabetes on fibrosis, further confirming the need for adjustment. The over-adjusted model tends to underestimate the CDE of diabetes, rendering it nearly nonsignificant. This difference in point estimates can be explained by adjustment on potential colliders. The wider confidence interval compared to our model can be attributed to two factors: (i) fewer data points are available in this estimation because it includes more variables, resulting in more missing data, and (ii) the causal adjustment set used in our model is optimal in the sense that it is proven to minimize estimation variance. For all these reasons, the causal discovery based estimation appears to be the most precise and reliable from a causal perspective.

**Figure 4.**
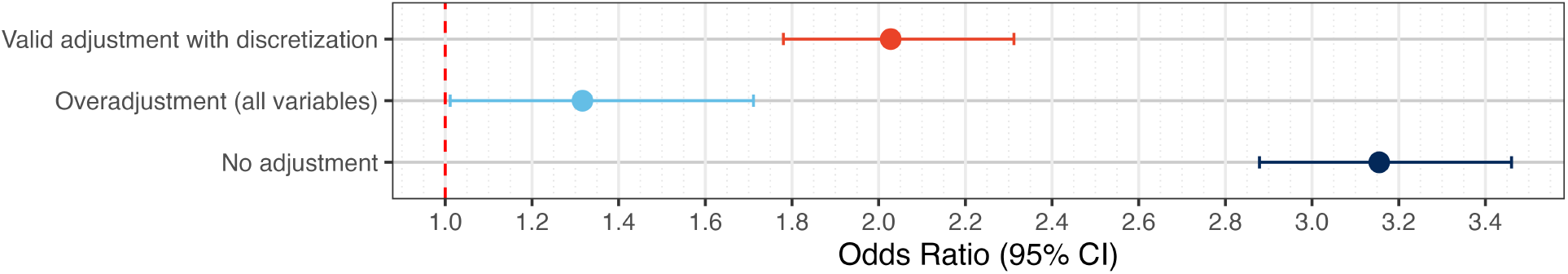
Estimated odds ratios for diabetes on severe fibrosis using the causal adjustment set, comparing discretized vs. original data, a naive unadjusted model, and an over-adjusted model.

## 4 DISCUSSION

This paper demonstrated the use of local causal discovery to estimate the CDE of diabetes on severe liver fibrosis in patients with chronic viral hepatitis. Traditional causal inference approaches often rely on expert-specified DAGs, which may be incomplete or partially unknown in complex epidemiological settings. Local causal discovery, by focusing only on the relevant portion of the graph necessary for a specific causal question, allows for data-driven identification of valid adjustment sets, reducing the assumptions and computational burden typically required by global causal discovery methods.

The application to the *ANRS CO22 HEPATHER* cohort to estimate the CDE of diabetes on severe liver fibrosis among patients with chronic hepatitis B, C, or B–C co-infection, showed that local discovery causal be useful to identify a correct adjustment set. Among 19 candidate variables, the algorithm identified 7 adjustment variables. Logistic regression yielded a controlled direct effect of diabetes on severe fibrosis with an odds ratio of 2.03 (95% CI: 1.78–2.31), suggesting a moderate but statistically significant effect. This application confirms the practical feasibility of local causal discovery in a real-world epidemiological dataset with multiple covariates and potential mediators, especially when it is used with robustness checks, such as bootstrapping and sensitivity analyses, are used to assess the stability of the estimated effects and the influence of potential unmeasured confounding.

We used the LocPC-CDE algorithm due to its simplicity and its specific design for estimating the CDE, but there exist other local causal discovery algorithms [38, 39]. Similarly, we employed logistic regression, though alternative methods for CDE estimation (such as odds ratios, risk ratios, or risk differences) can be used. It is entirely feasible to adopt other discovery algorithms or estimation methods while leveraging the pipeline we introduced in this paper: data preprocessing (discretization), incorporation of background knowledge, bootstrapping, extraction of the adjustment set, estimation, and sensitivity analysis.

We emphasize that the same discretized data were used for all estimation models, ensuring consistency with the data employed in the causal discovery phase. While this choice is particularly relevant for the present study, it would be possible to use non-discretized data to potentially improve the accuracy of the CDE estimation model.

Nevertheless, some limitations warrant consideration. Interpretation of the results presented in this paper relies on several assumptions. First, causal sufficiency and local faithfulness are assumed, meaning that all relevant variables are included locally and observed dependencies reflect true causal relationships. Violations of these assumptions, possible in biological systems (about violation of assumptions underlying causal discovery, see [40, 41, 8, 42]), may lead to missing or spurious edges and affect the identifiability of the causal effect. The sensitivity analysis mitigates concerns about a potential violation of causal sufficiency between diabetes and fibrosis: if an unmeasured confounder exists, it would need to have a strong effect to fully negate the significantly positive association observed. Second, data are assumed to be missing completely at random (MCAR); if missingness depends on measured factors, spurious associations may bias the estimated effect [31]. The MCAR assumption can sometimes be relaxed by performing global causal discovery [31], but this comes at a computational cost (in particular for a bootstrap-based approach as considered here), and global causal discovery generally performs worse than local causal discovery for the task of CDE identification [9]. Moreover, as in standard causal inference, we assume an underlying acyclic causal structure; feedback loops or cycles in biological processes could limit the validity of the conclusions.

Another important limitation of this paper is that we use a local causal discovery approach to select the relevant set of parents and then proceed to estimate causal effects using the same data. This practice compromises the coverage guarantees for the confidence intervals of the estimated causal effect [43]. In our study, since we used a test deletion strategy, the data used for causal discovery differs with the data used for the causal effect estimation. While this approach is likely to mitigate the issue, we cannot formally guarantee that it fully resolves it.

In conclusion, this study highlights that local causal discovery can serve as a valuable tool for causal inference in epidemiology, particularly when its limitations are carefully considered and appropriate robustness checks are applied. We emphasize that these methods are not intended to replace traditional approaches or expert knowledge, but rather to complement them, offering actionable insights for public health and clinical decision-making in situations where conventional methods alone may be insufficient.

## Data Availability

In regards to data availability, data from the study are protected under the protection of health data regulation set by the French National Commission on Informatics and Liberty (Commission Nationale de l'Informatique et des Libertés, CNIL). The data can be made available upon reasonable request to FC, after a consultation with the steering committee of the ANRS CO22 HEPATHER cohort study.

## Acknowledgements

The ANRS CO22 Hepather cohort received funding from Inserm-ANRS, Agence Nationale de la Recherche (ANR-11-EQPX-0021, ANR-19-COHO-0002), Direction Générale de la Santé (DGS), and Gilead, MSD, Abbvie, Janssen, BMS, Roche. This work was also supported by the

CIPHOD project (ANR-23-CPJ1-0212-01) and by funding from the French government managed by the National Research Agency (ANR) under the France 2030 program (ANR-23-IACL-0007). We would like to thank the study participants and the participating clinicians at each site.

## Notes

### Competing Interest Statement

Dr Bourliere reports consulting and lecturing fees from Gilead, AbbVie, GSK, Janssen, Roche and grant from Gilead and AbbVie
Other authors report no conflict of interest.

### Author Declarations

The ANRS CO22 HEPATHER protocol (ClinicalTrials.gov: NCT01953458) was developed in accordance with the Declaration of Helsinki and the French law for biomedical research. It was approved by the CPP Ile de France 3 ethics committee (Paris, France) and the French Regulatory Authority (ANSM). Written informed consent was obtained from each participant before enrollment.

### Summary of Updates

The causal discovery bootstrap was rerun with 100 iterations (previously 30), using a slightly revised implementation of the LocPC-CDE algorithm to ensure theoretical completeness. Specifically, symmetric conditional independence tests were incorporated to improve result consistency in line with recent theoretical developments on PC-style local causal discovery. The resulting inferred adjustment set differs marginally, with one variable (blood glucose) removed from the adjustment set. The overall findings of the paper remain consistent with the first version, with no impact on the methodological framework and no significant changes in the estimation results. In addition, the manuscript has been globally revised for improved clarity and conciseness.

